# Impaired systemic antibody response against gut microbiota pathobionts in critical illness and susceptibility to nosocomial infections

**DOI:** 10.1101/2025.07.08.25331079

**Authors:** Nicole A. Cho, Jared Schlechte, Ian-ling Yu, Ish Bains, Tanner Fahlman, Colin Mackenzie, Braedon McDonald

## Abstract

Critically ill patients in intensive care units (ICUs) experience high rates of nosocomial infections, commonly caused by translocation and dissemination of pathogenic microorganisms that colonize the intestinal tract (pathobionts). Multiple immune barriers protect the host against commensal and pathogenic colonizers, including a repertoire of circulating anti-commensal antibodies. The integrity of this systemic antibody-mediated defense system, its relationship with gut microbiota dysbiosis, and its impact on nosocomial infections in the ICU have not been explored. We observed markedly impaired plasma IgM and IgG reactivity against intestinal pathobionts such as *Escherichia coli*, *Klebsiella pneumoniae*, and *Enterococcus faecalis* in ICU patients compared to healthy volunteers. Reduced gut pathobiont antibody responses in ICU patients was associated with B cell lymphopenia, and patients with gut microbiota dysbiosis had reduced levels of natural antibody producing B1-like B cells. Reduced IgG against gut Gram-negative pathobionts was associated with an increased risk of nosocomial infection or death. These findings indicate that the systemic antibody barrier against microbiota pathobionts is compromised in critical illness and associated with increased risk of nosocomial infections, identifying a potential role for therapeutic antibody supplementation to prevent infections in the ICU.

## INTRODUCTION

Critically ill patients treated with life-support interventions in intensive care units (ICU) are at very high risk of nosocomial (hospital-acquired) infections, including ventilator-associated pneumonia (VAP), bloodstream infections (BSI), urinary tract infections (UTI), and *Clostridium difficile* infections (CDI), among others (1,2). Between 20-50% of mechanically ventilated ICU patients develop nosocomial infections (1,3,4), which is associated with a 2.5-fold increased risk of death compared to patients who do not develop nosocomial infections (5). This marked susceptibility to deadly infections in critical illness is often attributed to invasive interventions that breach normal barriers and create portals of entry for pathogens, such as intravascular devices, indwelling catheters, and endotracheal intubation. It is now well established that susceptibility to infections in critical illness is also the result of immune dysfunction causing impaired host defense against invading microbes, as well as pathological alterations to the host’s microbiome resulting in a reservoir of potential pathogens in the gut and other mucosal sites (6–10). As nosocomial infections remain a common and deadly threat to critically ill patients, there has been much attention to therapeutic and prophylactic strategies to reduce the burden of infectious complications in the ICU. Many of these interventions have been aimed at putative biological drivers including pathological microbiome alterations (digestive decontamination strategies, antimicrobial prophylaxis, probiotics), immune dysfunction, and care bundles (11–16). However, our incomplete understanding of the interplay between the gut microbiome and immune defenses in critical illness have likely contributed to the limited effectiveness of current strategies, yielding substantial opportunities to advance clinical care through improved understanding of microbiome-immune interactions.

The importance of microbiome dysbiosis in the pathogenesis of nosocomial infections is highlighted by the microbiology of infecting pathogens in the ICU. First, the majority of nosocomial infections are caused by organisms that are resident members of the microbiome, including Gram-negative pathobionts that reside in the gut (e.g. *Escherichia coli, Klebsiella spp*., *Enterobacter spp*., *Acinetobacter spp*.) and airway microbiota (e.g. *P. aeruginosa*), Gram-positive pathobionts of the gut (*Enterococcus spp.*) and skin and airways (*Staphylococcus spp.*), and fungal pathobionts (notably *Candida albicans)* (17,18). Secondly, these pathobionts are often residents of the human microbiota during homeostasis, and rarely cause serious infections in healthy people due to the presence of effective immune defense barriers that prevent translocation and dissemination of microbiota organisms in the body (19,20). This suggests that microbiota pathobionts act as “opportunistic” pathogens in the context of critical illness, possibly due to a breakdown of normal mechanisms that prevent translocation and dissemination of pathobionts during homeostasis (21). One such mechanisms is the production of antibodies in the blood directed against pathogenic microorganisms that colonize the gut and other mucosal surfaces (anti-pathobiont antibodies), which have been show in in animal models to serve as a crucial systemic immune barrier to pathobiont translocation and dissemination (22–25). However, the importance of anti-pathobiont antibody responses in humans is less understood.

Therefore, we conducted a study to test the hypothesis that healthy humans harbor anti-pathobiont antibodies in the blood, and that this mechanism of immune defense is impaired during critical illness in the setting of gut microbiota dysbiosis, thus contributing to the marked risk of nosocomial infections caused by gut pathobionts in critically ill patients. In this study, we quantified bloodstream antibody responses against 10 pathobionts representing the most commonly isolated Gram-negative, Gram-positive, and fungal pathogens in ICU nosocomial infections (26) in both healthy volunteers as well as critically ill patients at admission (day 1) and day 3 in the ICU. Anti-pathobiont antibody quantification was coupled with paired analysis of gut microbiota composition to investigate relationships between gut pathobiont (and commensal) colonization and circulating anti-pathobiont antibodies. Furthermore, we evaluated the relationship between anti-pathobiont antibody responses and nosocomial infection outcomes in ICU patients. We found that IgM and IgG responses against intestinal Gram-negative pathobionts were severely impaired in ICU patients, linked with dysregulated systemic B cell responses, and associated with an increased risk of nosocomial infections and death. Collectively, this study provides evidence that humans mount systemic, multi-isotype antibody responses against microbiome colonizers, and that during critical illness there is a severe impairment of systemic antibody reactivity to gut pathobionts that may contribute to the high susceptibility to nosocomial infections in the intensive care unit.

## RESULTS

### Participant population

To analyze anti-pathobiont antibody responses, their association with gut microbiota composition, and nosocomial infection outcomes, we prospectively collected blood and rectal swab samples at multiple timepoints during ICU admission from a cohort of 46 critically ill patients, compared to 28 healthy volunteers as controls. Demographic, clinical, and outcome data are displayed in Table 1. The current study represents a secondary analysis of participants in the MICRO-ICU study, from which additional gut microbiome and immunological analyses are available in a prior publication (6). The median age of ICU patient participants was 61 years, and 39.1% of patients were female. Inclusion and exclusion criteria (see details in methods) yielded a population of participants experiencing multi-system critical illness (median SOFA score of 8), all of whom were receiving mechanical ventilation support and enteral nutrition, and were free of overt confounders to immune and microbiome analyses (exclusion of immunocompromised individuals including those receiving immunomodulatory or immunosuppressive therapies, patients who were exposed to systemic antibiotics for >72h prior to enrollment, patients with discontinuous GI tract, or those hospitalized for >72h prior to enrolment). Enrolled participants had a range of admission diagnoses including sepsis (45.7%), trauma (26.1%), neurological emergencies (19.6%), and other (8.7%). Median duration of mechanical ventilation was 6 days, median ICU length of stay (LOS) was 7 days, and median duration of hospitalization was 17 days. Adverse outcomes were analyzed from ICU admission to day 30, including nosocomial infections that were diagnosed and treated in 56.5% of participants, and an overall 30-day mortality rate of 32.5% (Table 1).

**Table 1.**
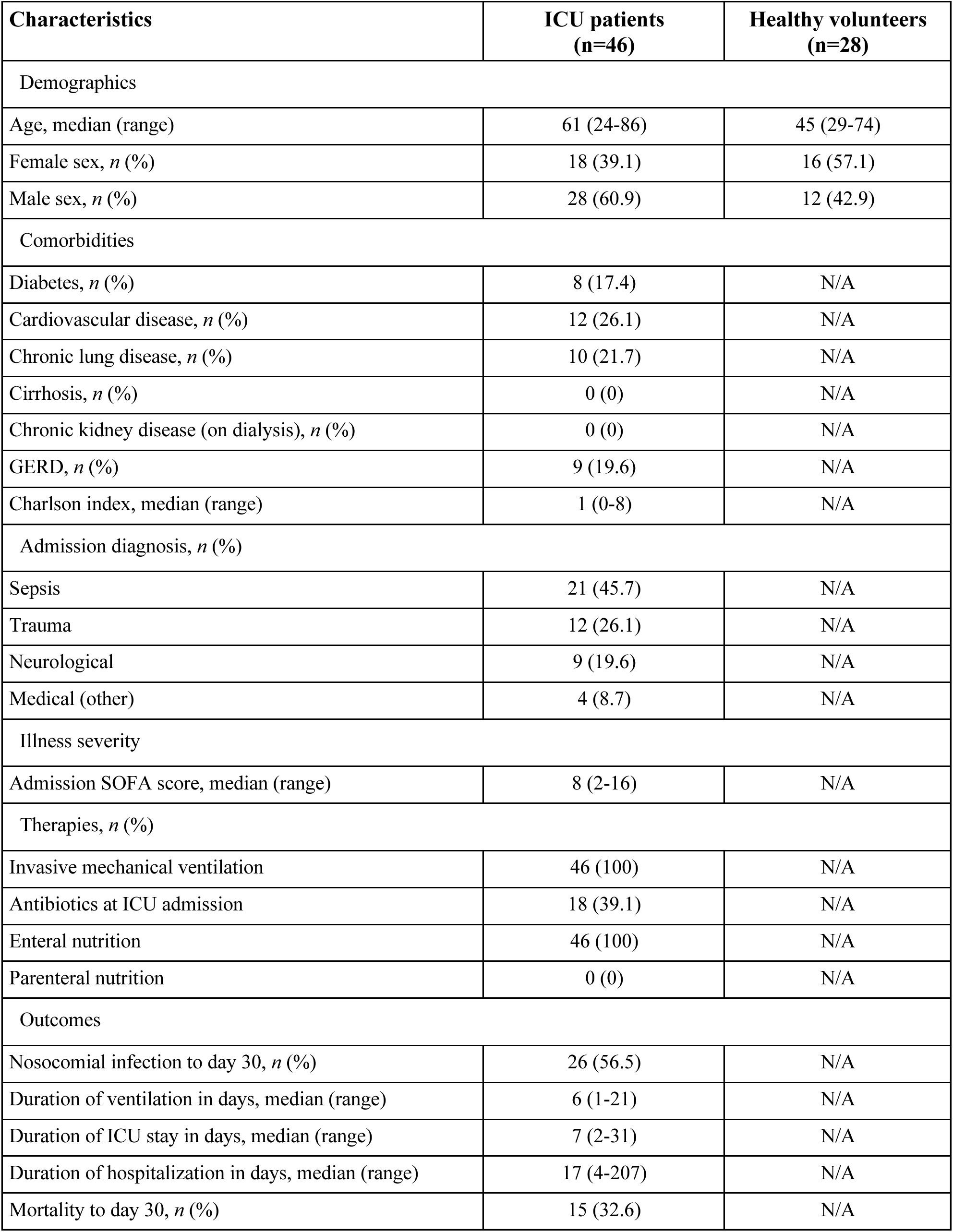
Characteristics of study participants.

### Critically ill patients demonstrate a sustained decrease in circulating IgM compared to healthy individuals

Sample collection schedule and experimental design are outlined in Figure 1A. Importantly, as the goal of this study was to investigate the contribution of anti-pathobiont antibody responses toward susceptibility to nosocomial infections (which are, by definition, hospital-acquired infections occurring >48h from admission), it was crucial to obtain samples prior to the onset of nosocomial infections. Therefore, blood and rectal swab samples were collected within 24h of ICU admission (day 1), and again on day 3 of admission, with all samples preceding the onset of nosocomial infections in this cohort (6).

**Figure 1.**
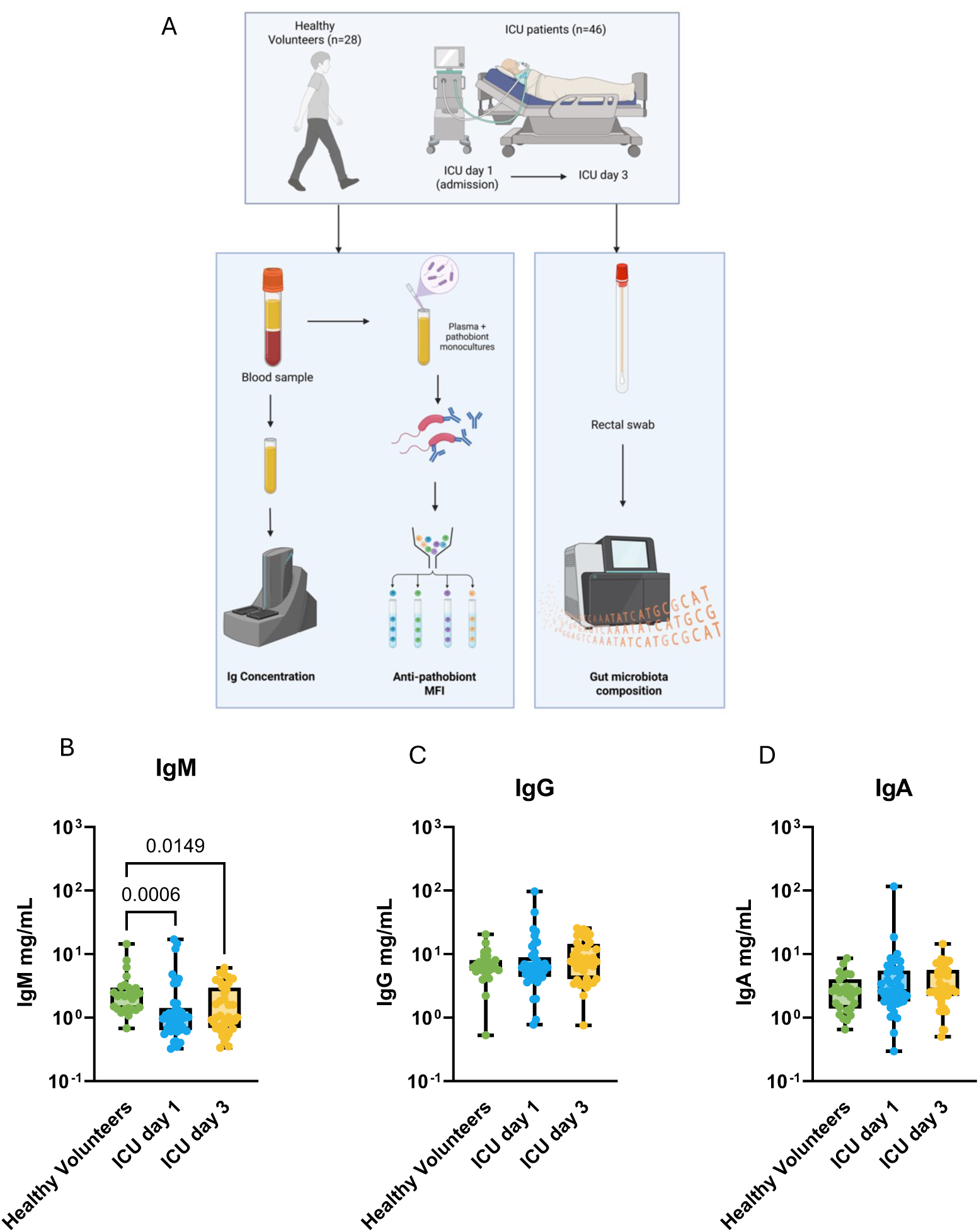
Systemic antibody responses in critically ill patients. (A) Flowchart of study design. (B-D) Quantitative analysis of (B) IgG, (C) IgM, and (D) IgA concentrations in the plasma of healthy volunteers (N=28) and critically ill patients (N=46) on day 1 and day 3 of ICU admission. Dots represent individual patients, central line indicates median, box shows interquartile range (IQR) and whiskers show range; analyzed by Kruskal-Wallis test with a post hoc Tukey’s test, significant p values as shown.

First, we quantified the concentration of total IgM, IgG, and IgA in the blood of ICU patients at admission (day 1) and compared it to concentrations observed in healthy volunteers. Consistent with the known circulating isotype distributions in humans, IgG concentrations were higher than IgM and IgA in the circulation in both healthy and critically ill patients (Figure 1B-D) (27). However, the median concentration of total IgM in ICU patients on day 1 of admission was significantly lower than healthy volunteer controls (Figure 1B), while no differences were observed in IgG nor IgA concentrations (Figure 1C-D). We next analyzed the temporal progression of total antibody concentrations in the circulation of ICU patients between admission and day 3, and observed stability of IgM, IgG, and IgA concentrations, with IgM concentrations on day 3 remaining significantly below levels seen in healthy volunteers (Figure 1B-D). Importantly, when stratified by admission diagnosis (sepsis, trauma, neurological, other), there were no differences in IgM, IgG, or IgA concentrations between these sub-groups of patients (Supplementary Figure 1).

### ICU patients have impaired antibody responses against gut microbiome pathobionts

To determine whether circulating antibodies against microbiome pathobionts are reduced in critical illness, we quantified IgG, IgM, and IgA binding in patient plasma samples to a library of 10 pathobionts representing the most common causative pathogens in ICU nosocomial infections using a flow cytometry (Table 2). In healthy volunteers, we observed the presence of anti-pathobiont reactivity for all isotypes, indicating that healthy adults possess circulating antibodies that are reactive to microbiome pathobionts (Supplementary Figure 2). Interestingly, gMFI of IgM and IgA reactivity was consistently higher than gMFI of IgG reactivity for all pathobionts tested (Supplementary Figure 2), despite lower total IgA and IgM concentrations compared to IgG (Figure 1 above).

**Table 2.** List of pathobiont microorganisms.

| **Organism** | **Strain** | **Source** | **Identifier** |
| --- | --- | --- | --- |
| *Staphylococcus aureus* | SH1000 | (Horsburgh et al., 2002) | N/A |
| *Staphylococcus epidermidis* | RP21 | Alberta Microbiota Repository (AMBR) | N/A |
| *Enterococcus faecalis* | KB1 | DSMZ | DSM 32036 |
| *Klebsiella aerogenes* | Eaer1 | Gift from Ian Lewis |  |
| *Klebsiella oxytoca* | K0x17 | Gift from Ian Lewis |  |
| *Klebsiella pneumoniae* | ST258 (isolate no. KpCG02) | (Chan et al., 2013) | N/A |
| *Enterobacter cloacae* | Eclo25 | Gift from Ian Lewis |  |
| *Escherichia coli* | ST131 | (McNally et al., 2016) | N/A |
| *Candida albicans* | Clinical isolate | (Bernardes et al., 2024) | N/A |
| *Pseudomonas aeruginosa* | RP9 | Alberta Microbiota Repository (AMBR) | N/A |

Gut pathobionts including Gram-negative bacteria (*E. coli*, *Klebsiella spp., Enterobacter spp.*) as well as Gram positive bacteria (*Enterococcus* spp.), have previously been reported to dominate the gut microbiome of many ICU and other acutely ill patients, which has been associated with immune dysfunction, as well as increased risk of nosocomial infections (6,28,29). These gut pathobionts are frequently causative pathogens in ICU nosocomial infections (1). Therefore, we analyzed circulating antibody reactivity against these common gut pathobionts in ICU patients compared to healthy volunteers (Figure 2). IgG reactivity against *E. coli*, *K. aerogenes,* and *E. faecalis* were all reduced in ICU patients at admission, with sustained reduction at day 3, compared to healthy volunteers (Figure 2A), whereas no differences were observed in IgG binding towards *K. oxytoca, K. pneumoniae,* and *E. cloacae* (Figure 2A). Summation of plasma IgG binding towards all Gram-negative gut pathobionts tested revealed an overall reduction in the cumulative anti-Gram negative pathobiont IgG reactivity at day 1 that was sustained to day 3 (Figure 2B). Compared to IgG, IgM binding to gut pathobionts was even more prominently impaired, with reduced plasma IgM binding to all gut pathobionts tested except *E. cloacae*, which demonstrated a non-significant trend towards reduction at admission and day 3 (Figure 2C). Overall anti-Gram negative pathobiont IgM reactivity was markedly lower at both day 1 and day 3 of ICU admission compared to levels observed in healthy volunteers (Figure 2D). Conversely, there were no differences in plasma IgA binding to gut pathobionts between ICU patients (either day 1 or 3) and healthy controls (Figure 2E-F).

**Figure 2.**
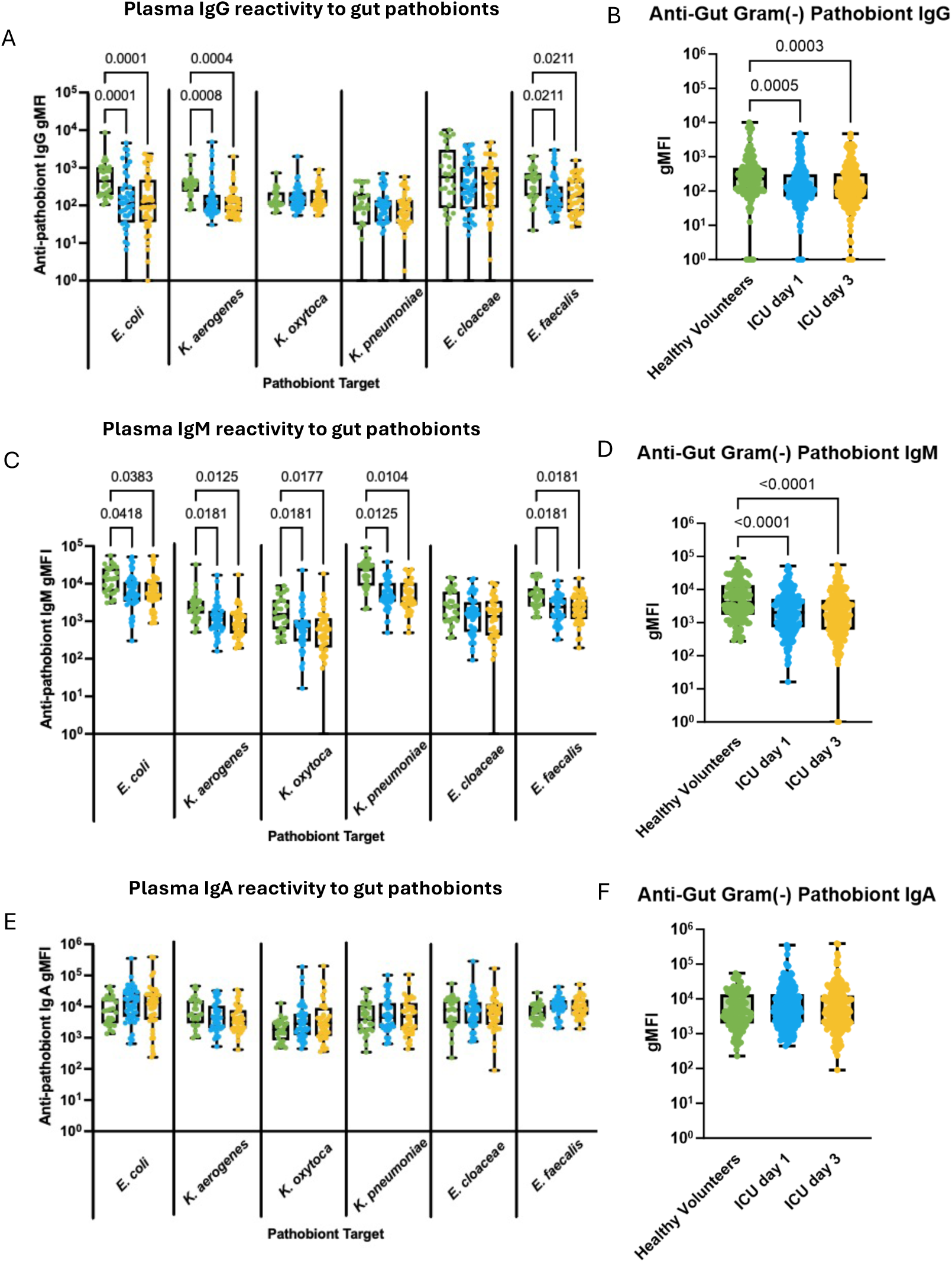
Systemic antibody responses against gut pathobionts in critically ill patients. (A) Flow cytometry was used to quantify plasma IgG binding to 6 individual gut pathobionts (expressed as geometric mean fluorescence intensity, gMFI) in healthy (N=26) and critically ill patients (N=46) on days 1 and 3 of ICU admission, and (B) shows pooled gMFI values for IgG binding to all gut Gram negative pathobionts together (*E. coli, K. pneumonia, K oxytoca, K. aerogenes, E. cloaceae*). The same analyses are shown for plasma IgM (C and D), and IgA (E and F) binding to gut pathobionts. Dots represent individual patients, central line indicates median, box shows interquartile range (IQR) and whiskers show range; analyzed by Kruskal-Wallis test with a post hoc Tukey’s test, significant p values as shown.

Of note, the large reduction of plasma IgM binding to gut pathobionts in ICU patients is unlikely to be a consequence of reduced total IgM concentrations in the blood. First, although total IgM concentrations are lower in ICU patients at admission and day 3 (Figure 1B above), this did not translate into reduced pathobiont-specific reactivity against all pathogens (as one would expect if it were solely due to lower IgM concentrations between groups), with no difference in reactivity towards *E. cloacae* between ICU and healthy participants. Next, IgG responses against gut pathobionts were also reduced, despite no difference in total IgG concentrations between ICU and healthy participants. Perhaps most importantly, as discussed further below, we observed increased IgM binding towards important skin pathobionts (see below). Collectively, these observations lead us to conclude that the reduced IgM and IgG binding to gut pathobionts observed in ICU patients is not attributed to lower immunoglobulin concentrations in the blood, but rather a consequence of altered gut pathobiont-specific humoral immunity.

Having identified important alterations in anti-gut pathobiont antibody responses in our diverse cohort of ICU patients, we sought to determine whether these antibody responses were associated with any important patient variables. Multiple linear regression analyses found that IgG, IgM, and IgA binding to gut pathobionts, as well as their total plasma concentrations were not significantly associated with age, sex, admission diagnosis, illness severity (SOFA score), burden of co-morbidities (Charlson index), nor duration of antibiotic exposure prior to sample collection (Table 3).

**Table 3:** Linear regression analysis of associations between patient characteristics, clinical measures, and antibiotics with antibody measurements.

| Variables | Models |  |  |  |  |  |
| --- | --- | --- | --- | --- | --- | --- |
|  | IgG anti-gut pathobiont gMFI | IgM anti-gut pathobiont gMFI | IgA anti-gut pathobiont gMFI | Total IgG concentration in plasma | Total IgM concentration in plasma | Total IgA concentration in plasma |
| Age | 0.274 (0.29) | 0.218 (0.34) | 0.043 (0.8) | 0.147 (0.45) | -0.0047(0.91) | 0.2391 (0.27) |
| Sex (Male) | 0.05 (0.53) | 0.041 (0.51) | 0.05 (0.3) | 5.684 (0.32) | 0.2767 (0.83) | 6.41 (0.31) |
| SOFA | -0.05 (0.71) | -0.041 (0.71) | -0.83 (0.5) | -0.078 (0.91) | 0.133 (0.45) | 0.207 (0.8) |
| Sepsis | 0.035 (0.6) | 0.043 (0.54) | 0.05 (0.46) | -3.187 (0.61) | -0.790 (0.59) | -2.944 (0.67) |
| Charlson Index | -0.03 (0.48) | -0.029 (0.43) | --0.01 (0.7) | -1.366 (0.68) | -0.599 (0.4) | -0.5323 (0.88) |
| Antibiotic days prior to sample | 0.007 (0.87) | 0.005 (0.92) | -0.015 (0.7) | 0.166 (0.96) | 0.2735 (0.78) | -1.704 (0.71) |

Data are Estimate (p value)

### ICU patients demonstrate variable systemic antibody responses towards extrta-intestinal pathobionts

Nosocomial infections in ICU patients are also commonly caused by pathobionts that reside outside the gut, in niches such as the skin (*S. aureus*, and coagulase-negative staphylococci like *S. epidermidis*, as well as the fungal pathobiont *Candida albicans*), as well as airway colonizers (*Pseudomonas aeruginosa*) (1,30,31). To determine whether systemic anti-pathobiont antibody responses differed against extra-intestinal pathobionts, we quantified plasma IgG, IgM, and IgA bindng towards *S. aureus*, *S. epidermidis*, *C. albicans*, and *P. aeruginosa*. Of note, we utilized a protein A (SpA)- and second immunoglobulin binding protein (Spi)- deficient strain of *S. aureus* (Δ*spA*Δ*sbi* SH1000) to avoid non-specific Fc binding of IgG in our assays. Contrary to what we observed towards gut pathobionts, there was a significant and sustained increase of plasma IgG, IgM, and IgA binding towards *S. aureus* in ICU patients on both days 1 and 3 compared to healthy controls (Figure 3A-C). Alternatively, antibody binding to the less pathogenic and less invasive skin pathobiont *S. epidermidis* was not different between ICU patients and healthy volunteers (Figure 3A-C). Like anti-*S. aureus* antibody responses, plasma reactivity towards the fungal pathobiont *C. albicans* was significantly increased in ICU patients for both IgG (Figure 3A) and IgA (Figure 3C), but not IgM (Figure 3B). For the airway pathobiont *P. aeruginosa*, decreased IgM reactivity (Figure 3B) was observed, without any change in IgG (Figure 3C) or IgA (Figure 3A) in ICU patients. Taken together, these data reveal an intriguing dichotomy between anti-pathobiont antibody responses in critically ill patients, with impaired responses towwards gut pathobionts (Figure 2) yet augmented responses towards skin pathobionts like *S. aureus* and *C. albicans* (Figure 3).

**Figure 3.**
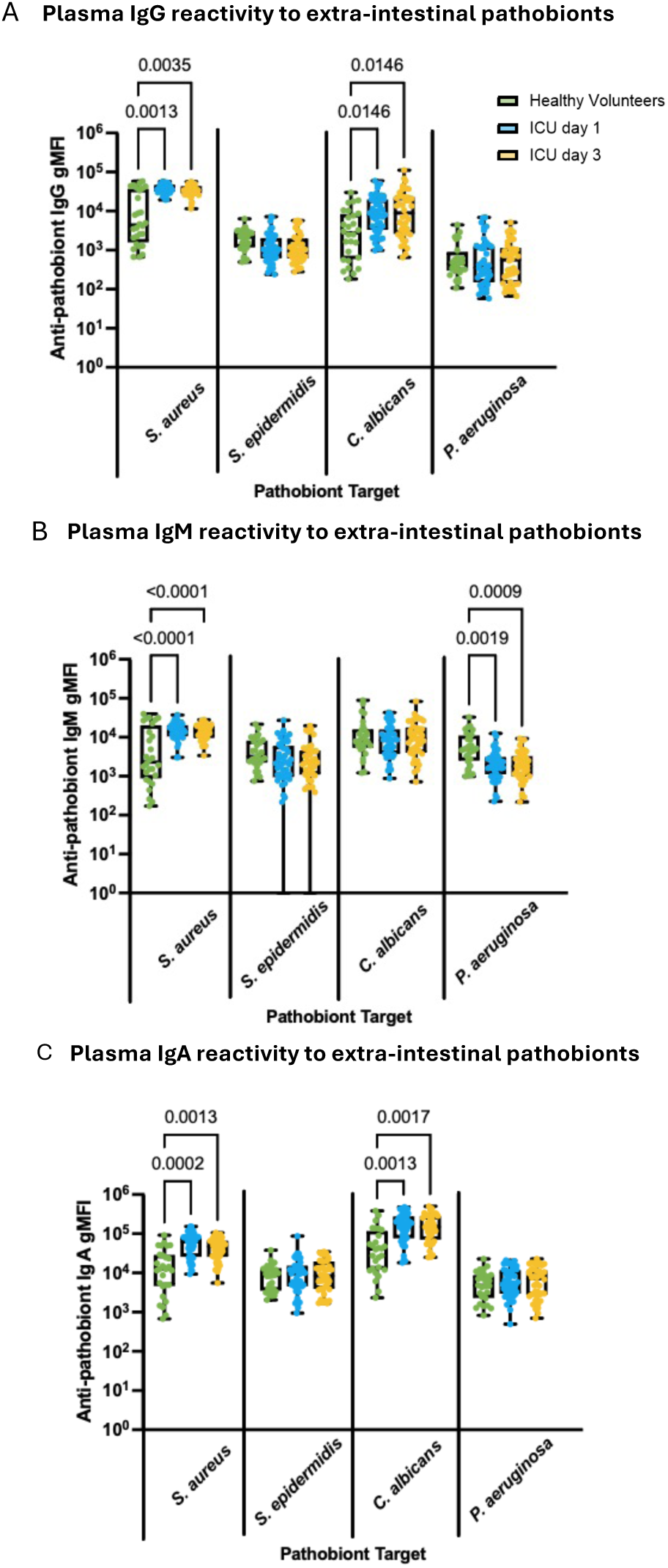
Systemic antibody responses against extra-intestinal pathobionts in critically ill patients. Flow cytometry was used to quantify plasma (A) IgG, (B) IgM, and (C) IgA binding to 4 extra-intestinal pathobionts (expressed as geometric mean fluorescence intensity, gMFI) in healthy (N=26) and critically ill patients (N=46) on days 1 and 3 of ICU admission. Dots represent individual patients, central line indicates median, box shows interquartile range (IQR) and whiskers show range; analyzed by Kruskal-Wallis test with a host hoc Tukey’s test, significant p values as shown.

### Circulating anti-pathobiont antibody responses and gut microbiota dysbiosis in critical illness

Hallmarks of gut microbiota dysbiosis in critical illness include reduced microbial biodiversity, and taxonomic alterations characterized by accumulation of pathobionts (most commonly Enterobacteriaceae such as *E. coli*, *Klebsiella* spp., *Enterobacter* spp., as well as Enterococcaceae *Enterococcus* spp.), as shown in previously published studies including prior gut microbiota profiling from the patients in this study (6). Given the prominent impairment of circulating antibody responses against gut pathobionts observed in ICU patients, we next investigated these humoral immune alterations were associated with composition of the fecal microbiota in each participant using matched samples. Correlation networks between anti-pathobiont antibody gMFI and the relative abundance of the top 20 taxa in the gut microbiota is shown for healthy volunteers in Figure 4A and 4C, and ICU patients in Figure 4B and 4D. Healthy volunteers were found to have strong and stereotyped correlations between plasma antibody reactivity against gut pathobionts and composition of the gut microbiota. In healthy volunteers there were strong positive correlations between plasma anti-pathobiont antibody quantities and intestinal abundance of *Enterobacteriaceae* and many anaerobic fermenter taxa, and negative correlations with Campylobacteriaceae, Peptostreptococcaceaea, and Veillonelaceae. In stark contrast, very few significant correlations were observed in ICU patients (Figure 4B and 4D). These data suggest that the relationships between gut microbiota composition and systemic anti-pathobiont antibody responses during health are lost in the context of gut microbiota dysbiosis in critical illness.

**Figure 4.**
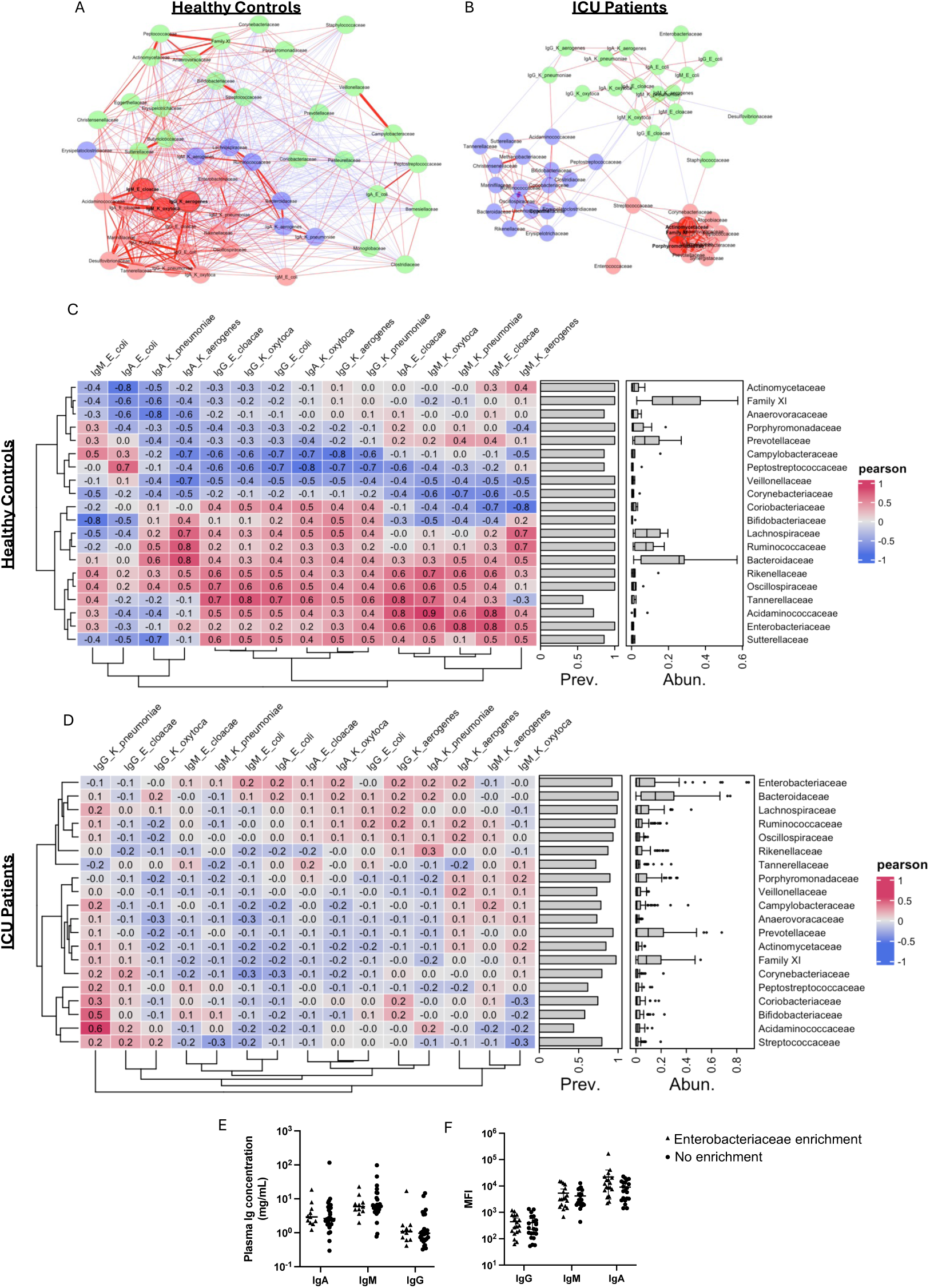
Relationships between plasma antibody binding to gut pathobionts and the composition of the gut microbiota in ICU patients and healthy volunteers. Pearson correlation networks of bacterial families (relative abundance) and plasma antibody binding to intestinal pathobionts (gMFI) in (A) healthy volunteer with antibody and gut microbiota datasets (N=7) and (B) ICU patients with matched antibody and gut microbiota datasets (N=42). Lines represent Pearson correlations between factors (red = positive correlation, blue = negative correlation) and line thickness representing strength of Pearson correlation. Nodes are coloured by cluster membership. (C-D) Heatmaps of Pearson correlation coefficients between the 20 most abundant bacterial families (relative abundance) and plasma antibody binding to gut pathobionts (gMFI) in (C) healthy volunteers (N=7) and (D) ICU Patients (N=42). Bacterial family prevalence and relative abundance are represented as bar graphs and box-and-whisker boxplots, respectively. (E) Concentrations of plasma IgG, IgM, and IgA, as well as (F) pooled gMFI of plasma antibody binding to gut pathobionts in ICU patients with and without gut microbiota Enterobacteriaceae enrichment. Dots represent individual patients, central line indicates median; analyzed by Kruskal-Wallis test with a host hoc Tukey’s test, no significant p values.

Previous work has demonstrated that patients with intestinal dysbiosis characterized by progressive Enterobacteriaceae enrichment have impaired innate immune function and increased risk of nosocomial infection or death (6). To investigate whether gut Enterobacteriaceae enrichment was also associated with alteration of humoral immunity against gut pathobionts, we compared antibody concentrations and anti-pathobiont reactivity between patients with and those without gut microbiota Enterobacteriaceae enrichment (as defined in Schlechte *et al.*(6)). However, we observed no difference in total IgG, IgM, or IgA concentrations, nor total anti-gut pathobiont IgG, IgM, nor IgA reactivity in plasma of ICU patients with gut Enterobacteriaceae enrichment versus those without (Figure 4E-F). Taken together, these data fail to demonstrate notable nor strong relationships between specific alterations of the gut microbiota during acute critical illness and the quantity of plasma IgG, IgM, or IgA reactivity against common gut pathobionts.

### Impaired anti-pathobiont antibody responses in critical illness are associated with dysregulated B cell responses

We quantified major populations of B cells in the circulation of ICU patients (day 1 and 3) and healthy volunteers using high-dimensional time-of-flight mass cytometry (CyTOF). As previously reported by us and others (6,32–34), we observed profound B lymphopenia in ICU patients at admission that was sustained to day 3 compared to healthy volunteers (Figure 5A). Analysis of B cell subsets (naïve, IgM+, class switched IgM-IgD-, and putative B1 (CD19+ CD1d+ CD43+, designated “putative” due to the lack of consensus surface marker definition for this rare population in human blood), revealed no difference in the number of naïve (Figure 5B), IgM+ (Figure 5C), nor putative B1 cells between ICU and healthy participants (Figure 5E). However, class switched (IgM-IgD-) antigen experienced B cells were significantly decreased in ICU patients on day 1, and this reduction was sustained at day 3 (Figure 5D).

**Figure 5.**
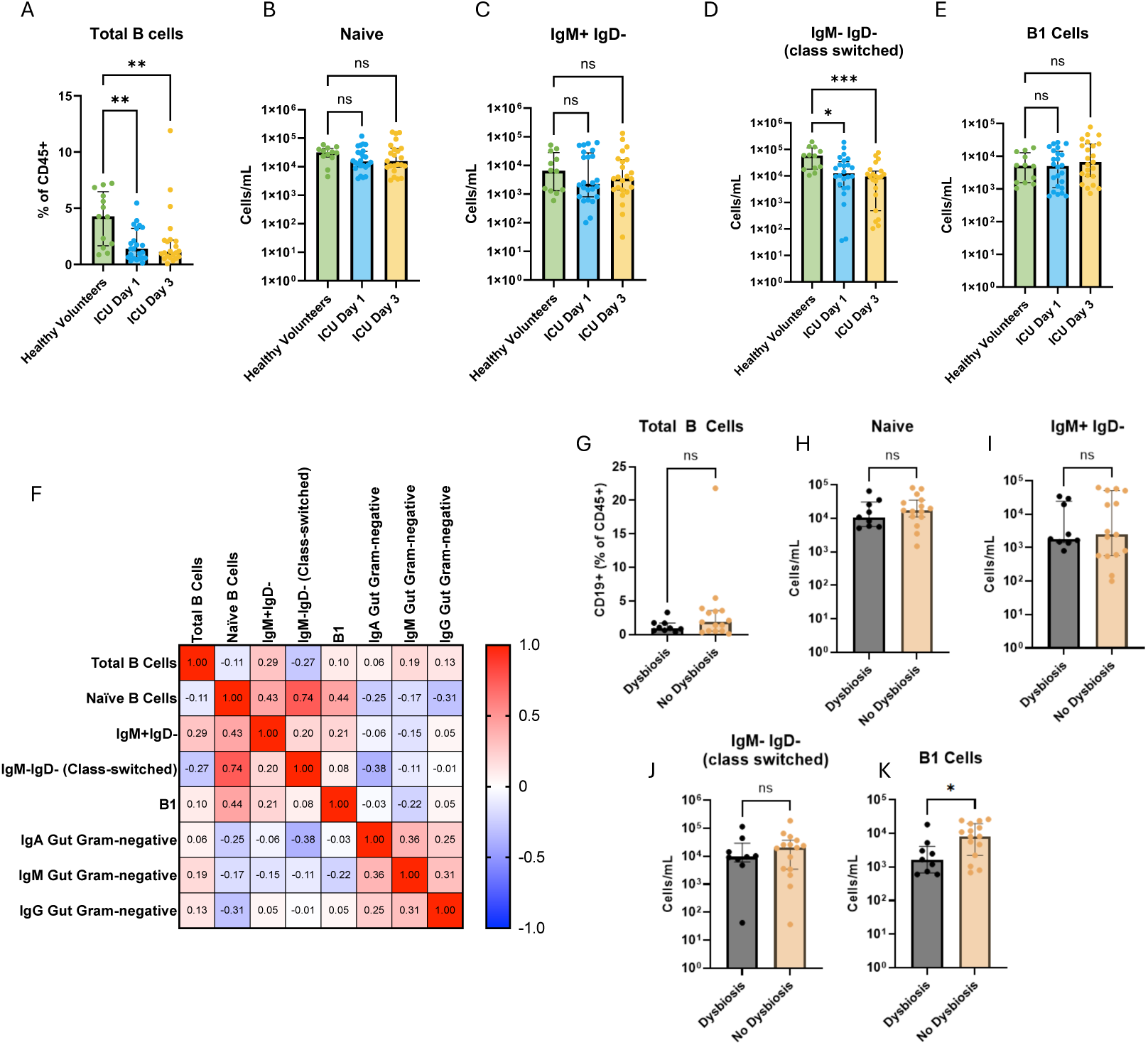
Systemic B cell responses and antibody reactivity against gut pathobionts in critically ill patients. Mass cytometry was used to quantify (A) total B cells (CD19+B220+ cells), (B) naïve B cells, (C) IgM+IgD-B cells, (D) Class-switched (IgM-IgD-) B cells, and (E) B1-like cells in blood from healthy volunteers (N=26) and ICU patients (N=46) on day 1 and 3 of ICU admission. (F) Spearman correlation matrix showing the relationships between quantities of B cell populations and anti-gut pathobiont IgG, IgA, and IgM reactivity (gMFI values) in the blood of ICU patients (numbers indicate Spearman’s correlation coefficient). (G-K) ICU patients were dichotomized into patients with gut microbiota dysbiosis (as defined in ref (6)) versus those without dysbiosis, and the quatities of the indicated B cell populations were compared. Dots represent individual patients, bars indicate median, error bars shows interquartile range (IQR); analyzed by Kruskal-Wallis test with a host hoc Tukey’s test (A-E) or Mann-Whitney U test (G-K). ***p<0.001, **p<0.01, *p<0.05, ns (non-significant, p>0.05).

Next, we interrogated the relationship between plasma IgM, IgG, and IgA binding towards gut Gram-negative pathobionts and circulating B cell populations, to understand the potential relationship between altered circulating B cells and impaired humoral response towards gut pathobionts. We observed negative correlations between IgM anti-gut pathobiont reactivity (gMFI) and the quantity of circulating B1, class-switched (IgM-IgD-), IgM+, and naïve B cells, while IgA and IgG against gut pathobionts were negatively correlated with class-switched and Naïve B cells (Figure 5F). Together, these data suggest that reduced anti-pathobiont antibody responses in ICU patients, particularly IgM responses, are associated with alterations of circulating B cell populations reflective of B cell dysregulation.

Prior literature, primarily from mouse models, have demonstrated that systemic anti-commensal antibody responses arise from microbiota-dependent regulation of multiple B cell compartments, including germinal centre reactions in secondary lymphoid organs as well as natural antibody production by B1 B cells. Therefore, we investigated whether patients with gut microbiota dysbiosis (Enterobacteriaceae enrichment, as defined in Schlechte *et al.*(6)) mounted differential B cell responses comparted to those without this signature of intestinal dysbiosis (no enrichment). Interestingly, while conventional B cell populations were equivalent between ICU patients with and without gut Enterobacteriaceae enrichment (Figure 5G-J), we observed significantly reduced quantities of circulating putative B1 cells in patient with Enterobacteriaceae dysbiosis (Figure 5K). These data raise the interesting possibility that gut microbiota composition may impact systemic B cell responses in critical illness specifically within the natural antibody producing B1 B cell compartment, which may contribute to the prominent dysregulation of circulating IgM responses in ICU patients.

### Impaired anti-gut pathobiont antibody responses in critically ill patients are associated with adverse outcomes including nosocomial infections and death

Having observed reduced plasma levels of IgM and IgG binding towards multiple gut pathobionts that commonly cause nosocomial infections in the ICU(6), we hypothesized that impairment of this humoral immune defense system may be associated with a higher risk of infections and related adverse outcomes including mortality. To test this hypothesis, we first stratified patients into 2 groups based on their anti-gut pathobiont antibody reactivity – those with anti-pathobiont antibody quantities (gMFI) above “normal” (median value of healthy volunteers) versus those with levels below normal. We found that patients with low anti-gut pathobiont IgG reactivity (gMFI below median of healthy controls) had significantly higher rates of nosocomial infections or death compared to patients with normal or elevated levels of anti-pathobiont IgG reactivity (Figure 6A). We saw no significant differences in rates of nosocomial infections or death between patients with high versus low IgM nor IgA reactivity (Figure 6B and 6C). Time-dependent analysis of nosocomial infection-free survival using maximal rank statistic determination showed that ICU patients with low levels of anti-*E. coli* IgM plasma reactivity (gMFI <4820) had significantly lower infection-free survival compared to ICU patients with higher plasma levels of anti-*E. coli* IgM (gMFI >/=4820) (Figure 6D). Collectively, these data suggest that reduced anti-gut pathobiont IgG responses, and reduced anti-*E. coli* IgM levels, are associated with an increased risk of nosocomial infections or death during critical illness.

**Figure 6.**
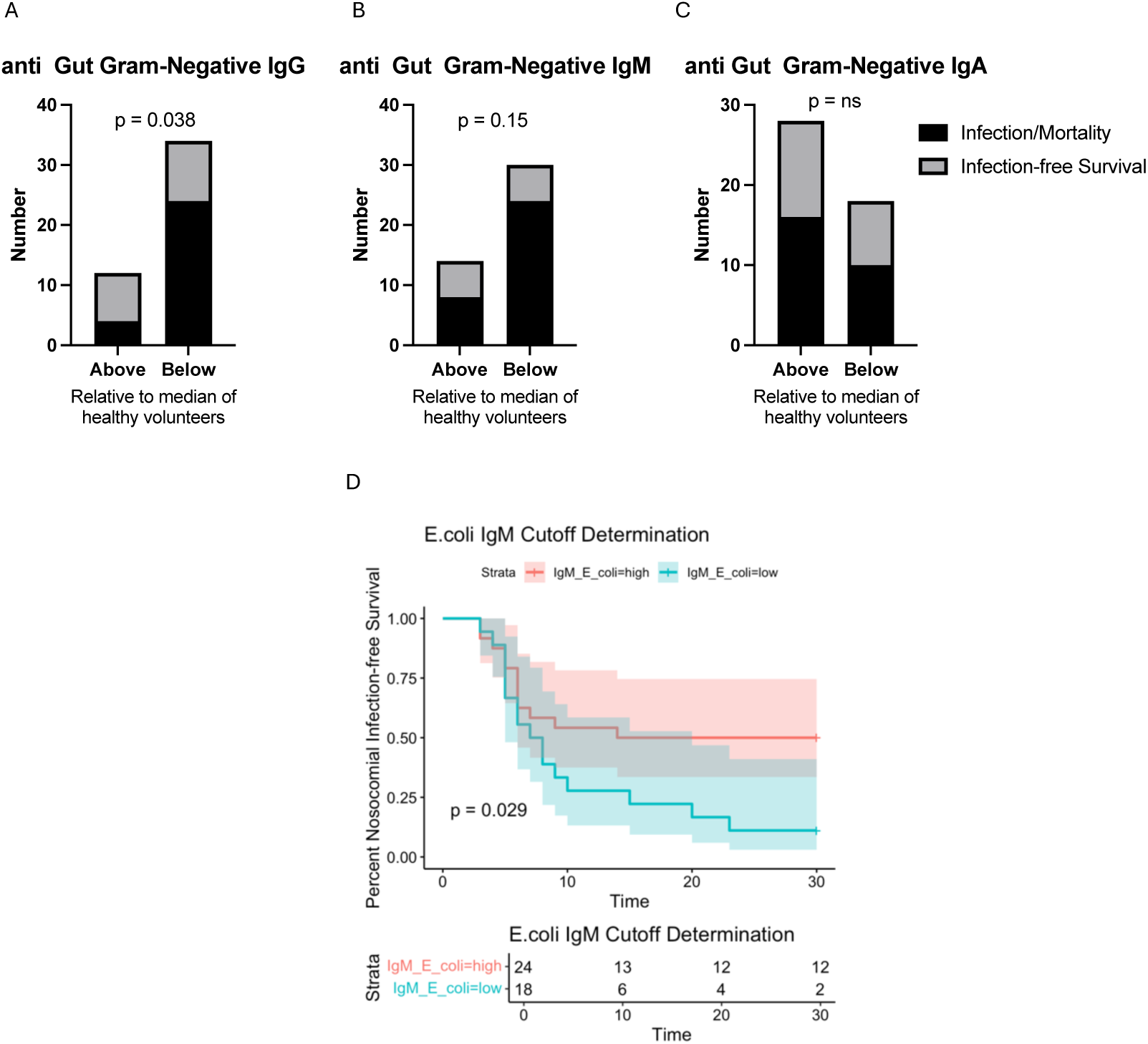
Association between systemic antibody responses against gut pathobionts and nosocomial infection-free survival. (A-C) Graphs show the number of ICU patients (N=46) who survived without nosocomial infections (black bars) versus those who developed nosocomial infections or died within 30 days of ICU admission (grey bars), comparing between patients with plasma antibody reactivity against gut Gram negative pathobionts (gMFI) above or below the “normal” level (median level of healthy volunteers); (A) IgG, (B) IgM, or (C) IgA). Data were analyzed using Fisher’s exact test, p values as shown. (D) Kaplan-Meyer curve displaying the proportion of ICU patients (N=46) with nosocomial infection-free survival to 30 days post ICU admission comparing individuals with high plasma anti-*E. coli* IgM (gMFI>/=4820) versus those with low plasmas anti-*E. coli* IgM (gMFI<4820). Analyzed by rank statistic, p=0.029.

### Impairment of anti-pathobiont antibody responses may disproportionately affect critically ill females

It is well established that many immunological diseases demonstrate sex-biased epidemiology, and in particular that B cell and antibody responses differ between males and females. In critical illness, the prevenance, severity, and outcomes of infections and sepsis differs by sex, but is overall understudied, underreported, and poorly understood. Therefore, we performed a sex stratified analysis of our data. We observed no differences in total IgG, IgM, or IgA concentration between males and females in healthy volunteers nor critically ill participants (Figure 7A-C). However, sex stratified analysis of anti-pathobiont antibody reactivity in plasma revealed that healthy females possessed higher anti-pathobiont IgG and IgM reactivity compared to healthy males, and that in critically ill patients, reduced anti-pathobiont IgG and IgM was seen only in females but not males (Figure 7D-E). This reduction in anti-pathobiont IgG and IgM was sustained from admission to day 3 in female patients (Figure 7D-E). Anti-gut Gram Negative IgA did not differ between female and male healthy volunteers, however critically ill females displayed higher anti-gut Gram Negative IgA compared to male patients on day 3 of ICU admission (Figure 7F). Additional sex-stratified analyses of individual pathobiont antibody binding are shown in Supplementary Figure 3. Collectively, these data suggest that impaired anti-pathobiont antibody responses during critical illness may disproportionately impact females as compared to males.

**Figure 7.**
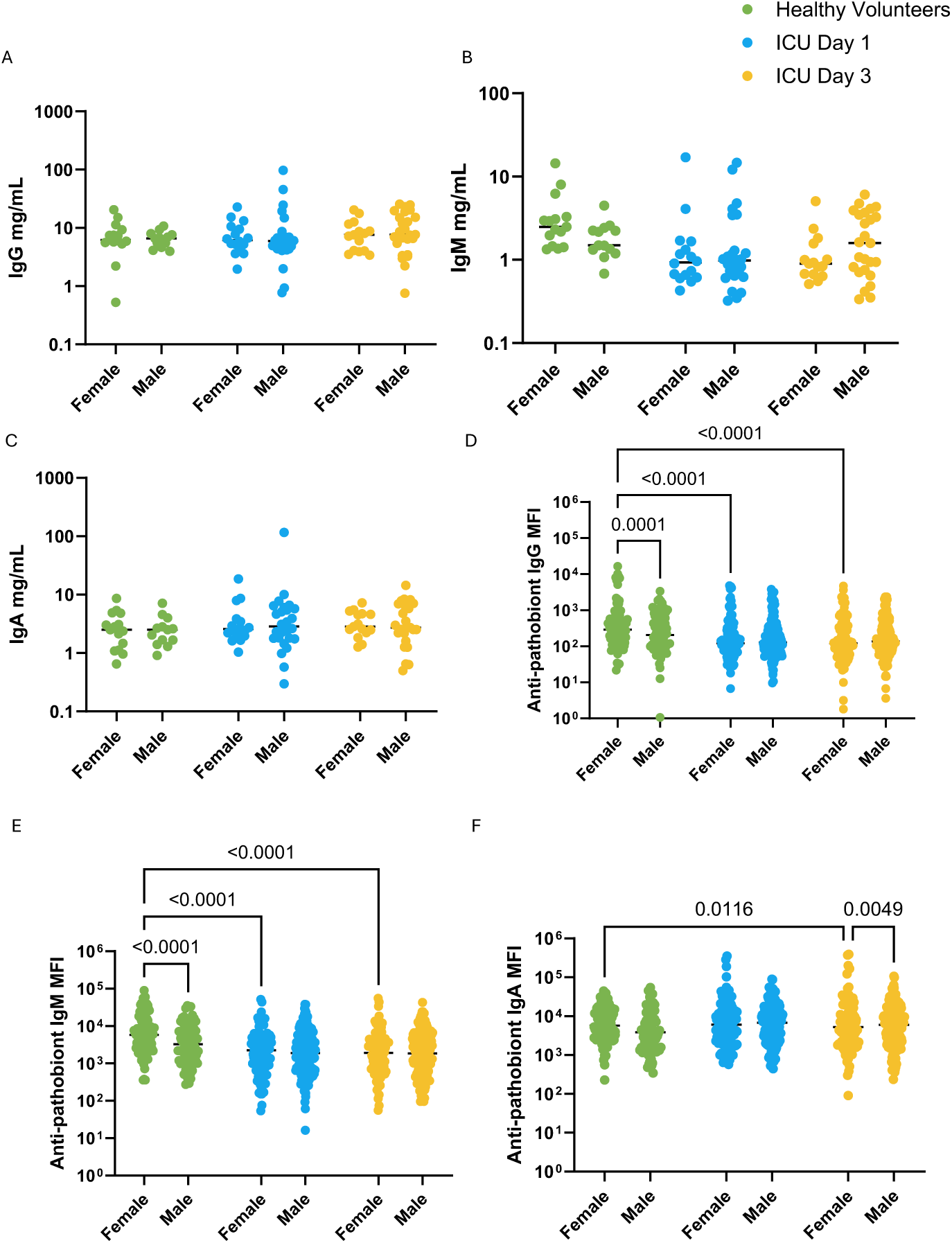
Sex disaggregated analysis of systemic antibody responses against intestinal pathobionts in critically ill patients. Flow cytometry was used to quantify plasma (A) IgG, (B) IgM, and (C) IgA binding to 4 extra-intestinal pathobionts (expressed as geometric mean fluorescence intensity, gMFI) in healthy (N=16 females, N=12 males) and critically ill patients (N=18 females, N=28 males) on days 1 and 3 of ICU admission. Dots represent individual patients, line indicates median; analyzed by Kruskal-Wallis test with a host hoc Tukey’s test, significant p values as shown.

## DISCUSSION

In this study, we report that humans mount systemic antibody responses against gut microbiome pathobionts that are impaired during acute critical illness. Prominent reductions of circulating IgM and IgG antibodies against intestinal pathobionts were observed in critically ill patients, and reduced IgM and IgG responses towards gut Gram negative pathobionts were associated with increased susceptibility to nosocomial infections. Matched analysis of fecal microbiota composition as well as systemic B cell responses revealed that gut microbiota dysbiosis, characterized by enrichment of Enterobacteriaceae pathobionts, was coupled with reduced circulating B1 B cells, a unique population of B cells well known to produce natural IgG and IgM antibodies, including against microbes that colonize mucosal surfaces.

Prior studies, primarily in mouse models, have established that systemic anti-microbiota antibodies in the bloodstream form a crucial barrier that protects against dissemination of microbes that translocate from mucosal surfaces into the blood (23,24,35). Indeed, acute and critically ill patients have been shown to display impaired gut barrier integrity, and translocation of gut microbes followed by dissemination through the bloodstream represents an important contributor to infections and end-organ dysfunction in these patient populations (10,36–38). Microbiological epidemiology of nosocomial infections in the intensive care setting consistently reports that more than half of ICU-acquired infections are caused by gut pathobionts, including Enterobacteriaceae and Enterococcaceae organisms (1). Therefore, our observation of significantly reduced circulating IgM and IgG reactivity towards Enterobacteriaceae and Enterococcaeae pathobionts may help to explain why ICU patients experience such a high risk of infections caused by these gut pathobionts, due to breakdown of this antibody-mediated barrier against the spread of microbiota organisms in the blood.

Dysbiosis of the gut microbiota has been linked with adverse outcomes including nosocomial infections in critically ill patients. In particular, expansion of Enterococaceae and Enterobacteriaceae in the gut of critically ill patients has been associated with infections and in-hospital mortality (6,39,40). Mechanistically, pathobiont expansion in the gut may potentiate infections during critical illness due to the establishment of a pathogen reservoir in the gut that can translocate in the setting of impaired mucosal barrier integrity, as well as adverse microbiota-immune interactions and impaired host defense (41). Indeed, Enterobacteriaceae expansion has been linked with innate immune dysfunction in critical illness, most notably neutrophil dysfunction (6). In the present study, matched analysis of individual patient circulating antibody, B cell, and gut microbiota data revealed that intestinal Enterobacteriaceae enrichment was associated with a reduction in putative B1 B cells. B1 cells are innate-like lymphocytes that are a main producers of natural antibodies, the repertoire of which is known to include IgM and IgG anti-commensal antibodies (42). Germ-free mice harbour reduced titres of microbiota-reactive serum IgM, indicating that the gut microbiota directs B1 cells to expand and secrete antibodies against mucosal colonizers (43). Interestingly, a recent study in critically ill patients as well as mice reported that depletion of B-1a lymphocytes and their natural antibodies resulted in profound susceptibility to infections by the environmental mucosal colonizer *Aspergillosis fumigatus* (44). This aligns with our observation of reduced B1-like cells in ICU patients with gut dysbiosis, and its association with increased risk of nosocomial infections. Similar to prior reports, we also observed total B lymphopenia with a prominent loss of class-switched (IgM-IgD-) B cells in critically ill patients compared to healthy controls. Therefore, the loss of anti-pathobiont IgM and IgG from the circulation of critically ill patients may be the result of both dysbiosis-related loss of natural antibody producing B1 cells, as well as the widespread reduction in classical B cell populations in critically ill patients.

Interestingly, we observed a dichotomy between circulating anti-pathobiont antibody responses directed against gut versus extra-intestinal pathobionts. While gut pathobiont-directed IgG responses were reduced in critically ill patients towards *E. coli*, *K. aerogenes*, and *E. faecalis*, we observed the opposite for skin-colonizing pathobionts *S. aureus*, and *C. albicans*, and no difference in IgG reactivity towards lung pathobiont *P. aeuruginosa*. Categorization of these organisms as skin and lung pathobionts is a generalization, as *S. aureus*, *C. albicans*, and *P. aeruginosa* are often multi-site colonizers including skin, respiratory tract, wounds, and even intestinal mucosa. The mechanisms driving these dichotomous response towards gut versus extra-intestinal pathobionts are unknown, but may relate to the unique microbiota changes that occur between the gut and extra-intestinal sites in critically ill patients. While our study did not observe a relationship between antibody responses towards extra-intestinal pathobionts and nosocomial infection, other studies of critically ill and hospitalized patients have suggested that reduced immunoglobulin responses against *S. aureus* (45,46) and *Candida spp.* (47) may contribute to infections caused by these organisms.

In addition to this interesting dichotomy in pathobiont reactivity, we also observed a striking difference between antibody isotype responses between IgG/M and IgA. IgA produced at mucosal sites is well known for its reactivity towards commensal organisms and central role in shaping the gut microbiota composition and protecting against pathobiont invasion (48). Interestingly, we observed no differences in systemic IgA reactivity towards any gut pathobionts between critically ill and healthy controls, while observing an increase in systemic IgA reactivity towards skin colonizers *S. aureus* and *C. albicans*. It will be of interest for future studies to investigate mucosal Ig responses in critically ill patients, to understand the relationship between microbiota dysbiosis and anti-pathobiont antibody responses at mucosal barrier sites versus the systemic circulation.

Our findings raise the interesting possibility that therapeutic supplementation of anti-pathobiont antibodies may protect against nosocomial infections in critical illness. Meta analysis of prior clinical trials of pooled donor immunoglobulin supplementation (IVIg) in sepsis have shown reduced mortality and length of hospitalization (49). IVIg contains a polyreactive repertoire of antibodies including against microbiota organisms, as well as many other proposed mechanisms of immunomodulatory activity, therefore the potential role of anti-commensal Igs within IVIg on outcomes in sepsis are unknown. However, given that our data showed prominently impaired IgM anti-pathobiont responses in ICU patients, it is notable that IgM-enriched formulations of IVIg have been reported to have the greatest efficacy in sepsis (49). Furthermore, monoclonal antibody therapies targeting selected pathobionts have also been investigated as prophylaxis against nosocomial infections by *S. aureus* and *P. aeruginosa* in the ICU (50–52). Our observations that systemic IgG responses against *S. aureus* were elevated, and IgG against *P. aeruginosa* was preserved, may help explain why clinical trials of monoclonal antibody therapies against these pathogens failed to prevent nosocomial infections in the ICU (50–52), as this approach may be redundant with high-levels of pre-existing endogenous antibodies towards these specific pathobionts. To date, the impact of therapeutic antibodies targetting gut pathobionts has not been addressed in humans. Taken together, our findings suggest that a precision approach to therapeutic antibody administration in which treatment is targeted at those patients with low circulating levels of endogenous anti-pathobiont antibodies may yield greatest efficacy to prevent nosocomial infections in the ICU.

Finally, we observed sex-specific anti-pathobiont responses in healthy volunteers as well as critically ill patients. Healthy females were found to express higher levels of anti-gut Gram-negative IgG and IgM reactivity compared to healthy males. Amongst critically ill patients, impairment of these antibody responses was greater in female participants compared to males. Prior studies have reported that healthy females have higher levels of circulating B cells (53), as well as more robust antibody responses to vaccines compared to males (54–56). Studies in both animal models as well as humans with severe infections and sepsis have reported sex-biased outcomes, with many studies finding higher incidence, severity, and mortality in males (57). Future studies of larger cohorts are needed to determine whether the observed sexual dimorphism of pathobiont-specific antibody responses are associated with the sex-biased clinical outcomes in critical illness.

This study has a number of limitations. First, the sample size was insufficient to explore the granular relationship between anti-pathobiont antibody responses and the specific pathogens causing nosocomial infections in ICU patients. This will be important to explore in larger future studies, as it is unknown whether our findings reflect dysregulation of antibody responses directed at specific pathobionts versus polyreactive antibody responses that broadly protect against diverse pathobionts. Next, our investigation was limited to a panel of 10 common pathobionts, and therefore it will be of interest for future studies to investigate antibody responses against additional pathobionts as well as commensal microbes. Dysregulation of systemic antibody responses against non-pathogenic commensals have been associated with inflammatory bowel disease pathogenesis (58). Furthermore, investigating additional anti-commensal antibody responses and its relationship with microbiota dysbiosis in critical illness may uncover further insight into the contribution of the intestinal dysbiosis towards nosocomial infections and outcomes in the ICU. Similarly, our study investigated only the bacterial microbiota and its relationship with anti-pathobiont antibody and B cell responses. Given that the fungal mycobiome in the human gut shapes systemic antifungal IgG binding, and individuals with systemic candidiasis mount a robust IgG response to Candida albicans (59), future studies should also look at how the mycobiome, and even virome, are linked to antibody production and patient outcomes.

In summary, this study reports that the systemic immune barrier formed by circulating anti-pathobiont antibodies is compromised in critically ill patients, and is associated with an increased risk of nosocomial infections. Our findings further clarify why critically ill patients suffer high rates of nosocomial infections caused by mucosal pathobionts, and suggest that a precision approach to therapeutic antibody supplementation in patients with low endogenous anti-pathobiont antibody reactivity may represent a strategy to prevent infections in the ICU.

## METHODS

### Sex as a biological variable

Data from both male and female patients and healthy volunteers were collected and analyzed.

### Study design and participants

This study was approved by the health research ethics boards of the University of Calgary and Alberta Health Services (REB18-1294). Male and female participants were included in this study, and sex as a biological variable was considered. Enrollment occurred between July 23, 2019 and July 20, 2021, with substantial delays and disruptions in enrollment due to the COVID-19 pandemic between March 2020 and April 2021. Written informed consent was obtained from all study participants or appropriate surrogate decision makers for patients who were unable to provide consent due to incapacitating illness. Patients admitted to the medical, surgical, neurological and trauma ICUs at the Foothills Medical Center in Calgary were screened for the following inclusion criteria (adapted from elsewhere(12)): adult (>18 years of age) with an index admission to ICU, requiring mechanical ventilation, expected to require continuous mechanical ventilation for >72 h as judged by the treating ICU specialist. Patients were excluded if they had a pre-existing immunocompromised state (systemic immunomodulatory therapy, chemotherapy, HIV infection or other congenital or acquired immunodeficiency), had been hospitalized >48 h before ICU admission in the previous 3 months, had received systemic antimicrobial therapy in the previous 3 months, had inflammatory bowel disease or active GI malignancy, previous surgery leaving a discontinuous GI tract, pregnancy, goals of care that excluded life-support interventions or moribund patients not expected to survive >72 h. At the onset of the COVID-19 pandemic, the study team added SARS-CoV-2 infection as an exclusion criterion and therefore no patients with COVID-19 were included in this study.

Samples were collected from ICU patients (N=46) on day 1 of ICU admission and again from survivors who remained in the ICU on day 3. For reference comparison, plasma samples were obtained from healthy volunteers (N=28). Population information detailed in Table 1.

### Plasma isotyping

The MSD Multi-Array Assay System (Human/NHP Isotyping Kit) Multiplex kit was used to determine concentrations of IgA, IgG, and IgM according to the manufacturer’s instructions

### Bacterial and fungal FACS

Bacterial and fungal strains used in this study are listed in Table 2. Subcultures were made from overnight monocultures of each listed pathobiont and diluted to 2×10^7^ CFU/mL in FACs wash buffer. Serum was incubated at 56°C for 30 minutes, and centrifuged at 4,000g for 10 minutes to inactivate complement. 100ul of subcultures were plated in a 96 well V-bottom plate, and 5ul of serum was added to each well to incubate for 1 hour at room temperature, shaking at 300rpm. The wells were washed twice with FACs wash buffer, then cells were resuspended in 50ul FACs wash buffer containing 1:50 IgA-PE (Miltenyi Biotec), 1:50 IgG-AF647 (Biolegend), and 1:50 IgM-PE/Cy7 (Biolegend) and incubated overnight at 4°C. The next day, wells were washed with PBS and resuspended in 100ul of 4% PFA/PBS for 15 minutes at room temperature to fix cells, then washed again with PBS. Cells were resuspended in 100ul of 1:200 DAPI/PBS for bacteria or 1:2000 CFW/PBS for *C. albicans* cultures and acquired on CytoFlex LX (Beckman Coulter).

### 16S rRNA gene amplification and sequencing, sequence data processing and analysis

This study contains a secondary analysis from a previously published dataset, where methods are available(6). Briefly, rectal swabs were collected and stored in sterile tubes at −80 °C, and DNA was isolated using the DNeasy PowerSoil kit (QIAGEN) according to the manufacturer’s protocol. PCR amplification of the 16S V4 region was performed and sequenced using an Illumina MiSeq platform. De-multiplexed Illumina MiSeq paired-end reads (FASTQ) were processed in R v4.1.2 following the DADA2 pipeline v.1.14. Taxonomy of unique amplicon sequence variants (ASVs) was assigned in DADA2 using the SILVA v.138.1 database. ASVs and sample data were combined using the Phyloseq package v.1.38.0 for further downstream analysis.

### Time of Flight Mass Cytometry

Single cell time-of-flight mass cytometry (CyTOF) was acquired as previously described(6). Whole blood was cryopreserved in PROT1 proteomic stabilizer (SmartTube Inc.) at −80°C following sample collection. For acquisition, samples were thawed and reb blood cells were lysed using Thaw-Lyse buffer (SmartTube Inc.), and the single cell suspension was then passed through a 70 uM cell strainer. Cells were then fixed and permeabilized and subsequently barcoded with a unique 3 metal barcode using the Cell-ID 20-Plex Pd Barcoding Kit following manufacturers protocol (Standard Biotools). Following barcoding, samples were pooled and then stained with a cocktail of metal-labeled antibodies, then fixed overnight with DNA intercalator - Cell-ID™ Intercalator-Ir – and the following day the pooled samples were acquired on a Helios II. The acquired data was de-barcoded and analyzed using Cytobank (Beckman Coulter).

### Statistical analysis

Antibody data are presented as median and IQR. Normality was assessed using the Shapiro-Wilk test in GraphPad Prism. Differences between groups were assessed using Kruskal-Wallis test with Tukey’s post hoc test for multiple comparisons. Multiple linear regressions and correlations were performed in GraphPad Prism. Results were considered significant at *p*<0.05. E. coli IgM gMFI cutoff and survival analysis was performed in R using the survminer R package.

Microbiome correlation networks with antibody concentrations and gMFI were generated using the NetCoMi R package (60). Microbiome data was filtered using a 25% prevalence filter and the remaining taxa were aggregated to the family level and log transformed. Antibody concentration and MFI data was log transformed, and a Pearson correlation was calculated between microbiome data and antibody data.

## Data availability

Microbiome DNA sequence datasets available in the NCBI Sequence Read Archive under BioProject ID PRJNA851469. Additional datasets and code are available from the corresponding author upon reasonable request.

## AUTHOR CONTRIBUTIONS

B.M. conceived the study, designed experiments, secured funding and provided overall supervision. I.B and B.M consented and enrolled participants, and I.Y. I.B., and B.M. collected, processed and stored patient samples. I.Y. and T.F performed antibody analysis experiments. N.A.C., J.S., C.M., and B.M. analyzed data and prepared figures. N.A.C. and B.M. prepared the manuscript. All authors edited the manuscript.

## ACKNOWLEDGMENTS

The authors thank the patients and families who contributed to this study. The authors would also like to thank Yiping Liu and the Flow Cytometry Core Facility, Cumming School of Medicine, for assistance with acquiring flow cytometry sample data. This work was supported by operating grants from the Canadian Institutes of Health Research (CIHR) project grant (grant no. 173296), and an CIHR Early Career Investigator Award in Infections and Immunity (grant no. 170746), Canadian Foundation for Innovation JR Evans Leaders Fund Grant (grant no. 40697), all to B.M. NAC is supported by an Eye’s High Postdoctoral Fellowship from the University of Calgary and a CIHR Postdoctoral Fellowship (FRN 194087). JS is supported by a CIHR Doctoral Studentship Award. CM is supported by a CIHR Postdoctoral Fellowship (FRN 201059).

## SUPPLEMENTARY FIGURES 1-3

**Supplementary Figure 1.**
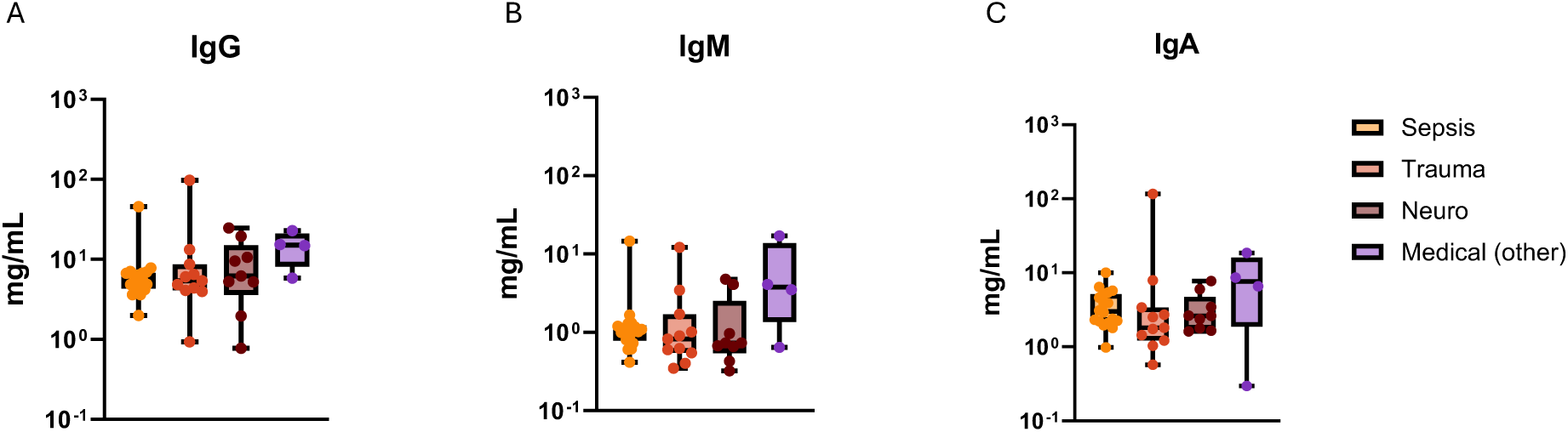
Systemic antibody levels in ICU patients stratified by admission diagnosis. Quantitative analysis of (B) IgG, (C) IgM, and (D) IgA concentrations in the plasma of critically ill ICU patients (N=46) on day 1 stratified by admission diagnosis. Dots represent individual patients, central line indicates median, box shows interquartile range (IQR) and whiskers show range; analyzed by Kruskal-Wallis test with a post hoc Tukey’s test, p values non-significant (p>0.05).

**Supplementary Figure 2.**
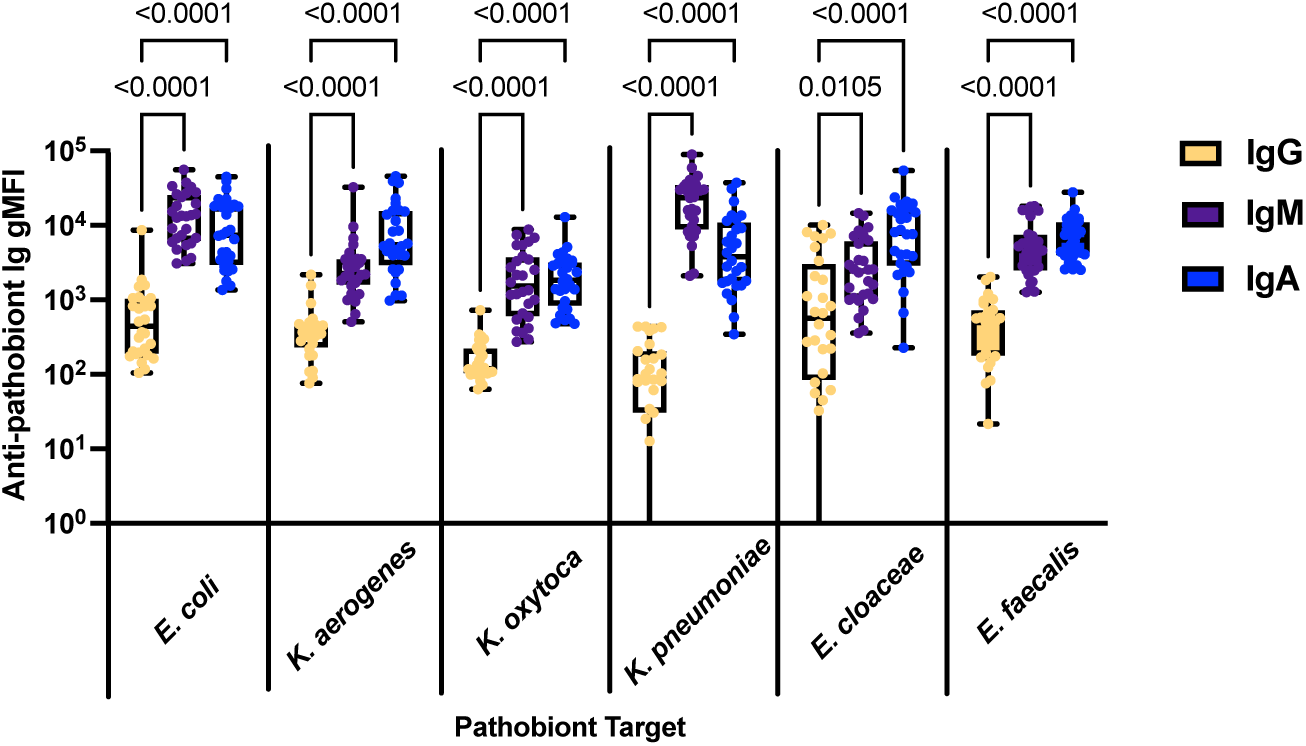
Quantitative comparison of plasma IgG, IgM, and IgA binding to gut pathobionts in healthy volunteers. Flow cytometry was used to quantify plasma IgG binding to 6 individual gut pathobionts (expressed as geometric mean fluorescence intensity, gMFI) in healthy volunteers (N=26). Dots represent individual patients, central line indicates median, box shows interquartile range (IQR) and whiskers show range; analyzed by pathobiont using Kruskal-Wallis test with a post hoc Tukey’s test, significant p values as shown.

**Supplementary Figure 3.**
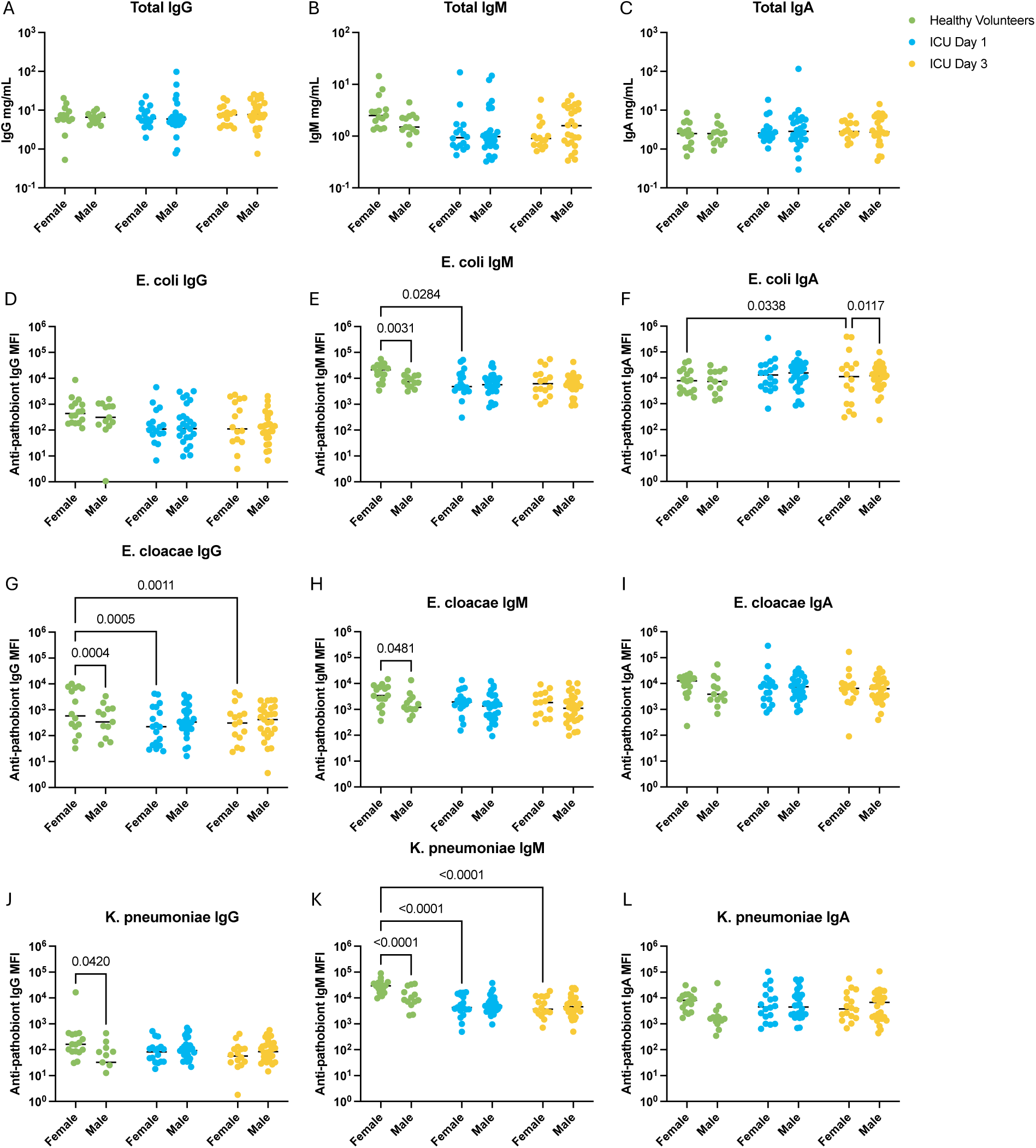
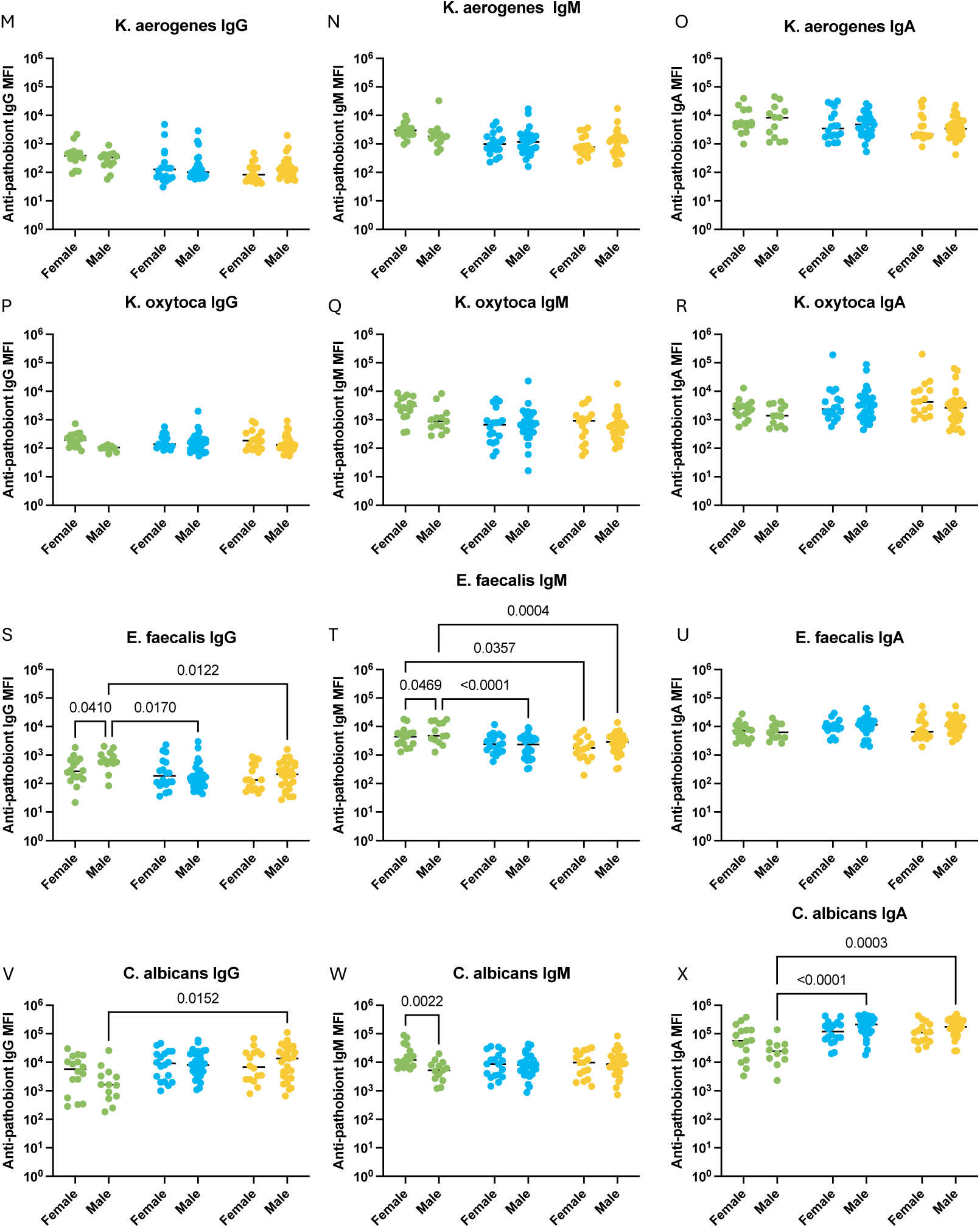
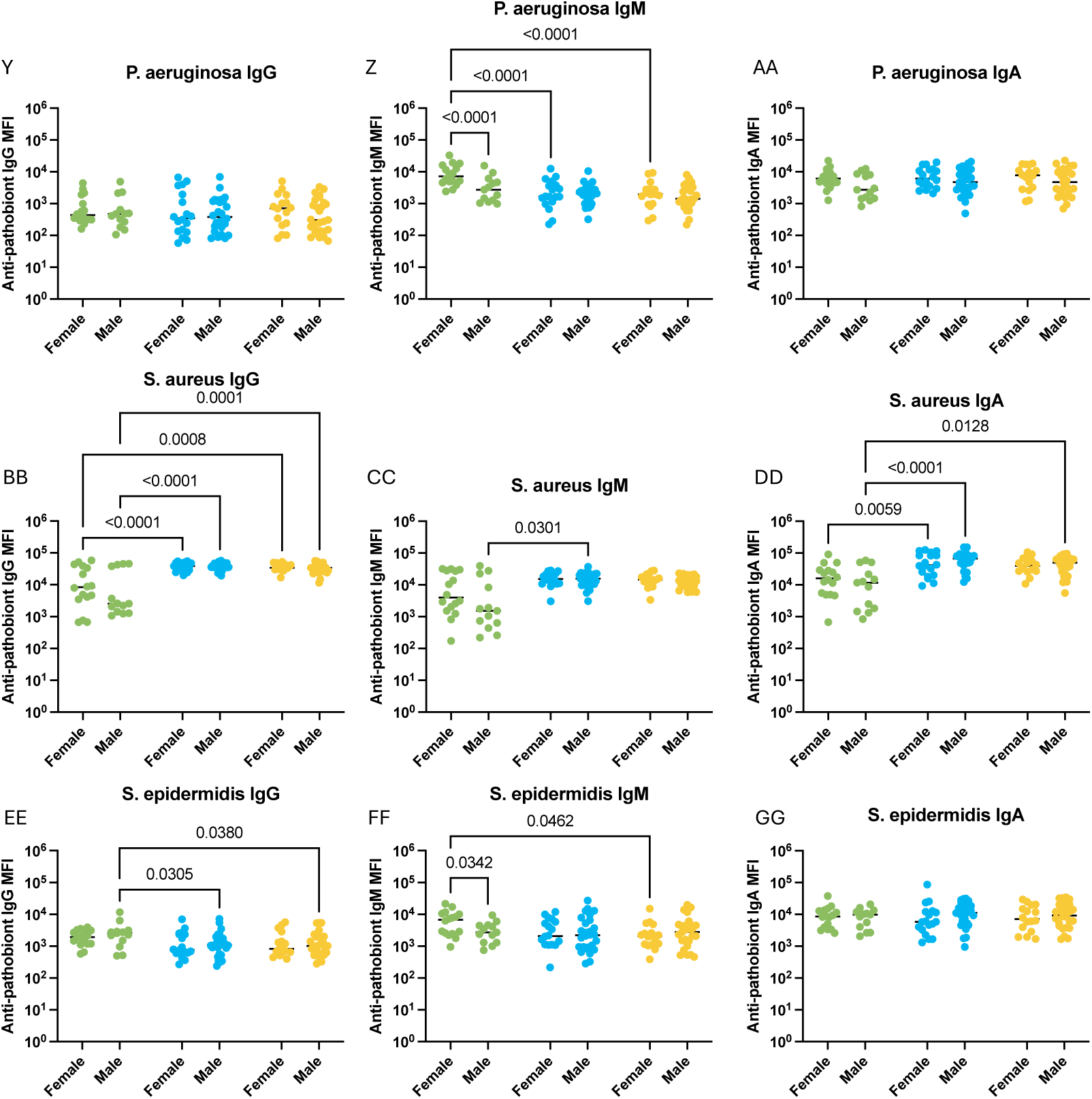
Sex disaggregated analysis of systemic antibody responses against pathobionts in critically ill patients. Flow cytometry was used to quantify plasma (A) IgG, (B) IgM, and (C) IgA binding to 10 pathobionts (expressed as geometric mean fluorescence intensity, gMFI) in healthy (N=16 females, N=12 males) and critically ill patients (N=18 females, N=28 males) on days 1 and 3 of ICU admission. Dots represent individual patients, line indicates median; analyzed by Kruskal-Wallis test with a host hoc Tukey’s test, significant p values as shown.

